# Clarifying the role of Cortico-Cortical and Amygdalo-Cortical brain dysconnectivity associated with Conduct Problems

**DOI:** 10.1101/2022.08.01.22278272

**Authors:** Jules R. Dugré, Stéphane Potvin

## Abstract

**Background:** Recent evidence suggests that adolescents exhibiting conduct problems (CP) may have disrupted brain connectivity at rest. However, these studies are generally characterized by small sample sizes and differ in terms of neuroimaging methodologies and chosen psychometric scales to assess CP. In parallel, evidence from genetic and structural imaging studies suggests that aggregating measures may increase generalizability and reproducibility in results. Our primary aim was to identify deficits in functional brain connectivity that were replicable across two distinct measures of CP.

**Methods:** In a large sample of adolescents (n=1416), we assessed the relationship between resting-state functional connectome (including the amygdala bilaterally) and two distinct measures of conduct problems. Positive and negative brain connectivity measures were derived from the intersection of both scales. The utility of these aggregated scores was assessed in comparison with variants of psychopathy and clinical diagnoses.

**Results:** Psychometrics scales assessing CP were significantly related to 231 & 269 disrupted functional connectivity. Only 21 brain connectivity were shared between the two scales (10 positively and 11 negatively associated with CP. These brain connectivity scores differed between adolescents with variants of psychopathy and healthy controls and were specifically associated with disruptive disorders, but not other pediatric psychiatric disorders.

**Conclusion:** The current study provides the evidence that different measures of CP may yield distinct results. Nonetheless, it also highlights that functional brain connectivity that intersected between the two scales may be robust and reliable neurobiological markers of severity of CP. Finally, brain connectivity scores may be generalizable to variants of psychopathy and specific to disruptive disorders.

## 1. INTRODUCTION

Conduct disorder (CD) is defined by serious and persistent patterns of behavior that violate the rights of others (i.e., aggressive, and rule-breaking behaviors) (1). It has been suggested that approximately 5% of children will display severe and persistent conduct problems (CP) and meet the criteria for CD (2, 3). These children are known to display high levels of comorbid psychopathologies such as callous-unemotional traits (up to 50%, (4), but also attention-deficit/hyperactivity (ADHD) symptoms, anxiety and mood problems (5-9). Interestingly, past studies have shown that individuals with CP may demonstrate a variety of neurobiological impairments. For instance, in a recent meta-analysis of functional neuroimaging studies, our research team observed that this population was characterized by abnormal brain activity during fMRI tasks involving negative emotions processing, social cognition and cognitive control (10). Indeed, the literature has extensively supported the role of the amygdala, medial and lateral prefrontal cortex, insula and cingulate cortex in our understanding of the neural correlates of CP during functional magnetic resonance imaging (fMRI) tasks (10-14). However, in comparison to other neuroimaging modalities, the neurobiological markers of functional connectivity at rest in CP remain understudied.

In the last decade, researchers have aimed to identify intrinsic functional networks which regroup reliable temporally correlated brain regions at rest. Indeed, these large-scale networks usually include the medial fronto-parietal (e.g. default-mode network [DMN]), occipital (e.g. medial and lateral visual), pericentral (e.g. sensorimotor, somatomotor [SomMot]), dorsal fronto-parietal (e.g. dorsal attention [DorsAttn]), lateral fronto-parietal (e.g. cognitive control), midcingulo-insular (e.g. salience [SAL], ventral attention [VentAttn], cingulo-opercular [CingOperc]) networks (15-18). Recently, resting-state functional connectivity has gained considerable attention in the investigation of the neurobiological mechanisms involved in antisocial behaviors. For example, in a recent meta-analysis of resting-state functional connectivity studies from our research team, we found that antisocial subjects exhibited prominent functional connectivity alterations in the DMN (i.e., ventro- and dorso-medial PFC and posterior cingulate cortex), DorsAttn (i.e., Frontal eye field) and VentAttn regions (i.e., anterior midcingulate cortex/pre-supplementary motor area) as well as in the amygdala (19). Using the Gordon Atlas from the ABCD study (n=9636), it has been shown that the severity of CP was associated with average within-connectivity in the DorsAttn, whereas reduced connectivity within the DMN was associated with callous-unemotional traits (20). Similarly, reduced PCC-vmPFC connectivity (within-DMN) was found when comparing inmates with and without psychopathic traits (21). However, some recent meta-analytic evidence suggests that dysconnectivity of the DMN may not be specific to CP but rather act as a transdiagnostic neurobiological markers (22). These results justify the need to search for specific neurobiological markers of CP.

Growing evidence suggests that antisocial behaviors may be mostly associated with impairments of between-rather than within-network connectivity. For example, in a large sample of adults (n=1003), some researchers have found that anger-aggression was mainly correlated with connectivity between the PCC (DMN) and visual, SomMot and VentAttn as well as between the FP and SomMot (23). Furthermore, dysconnectivity between VentAttn regions and DMN (24) and FP (25) as well as between the DorsAttn and DMN and VentAttn, were found to be associated antisocial behaviors (26). Furthermore, despite that the amygdala is not included in a particular brain network defined at the cortical level, some researchers found that subjects exhibiting antisocial behaviors demonstrated altered resting-state connectivity between the amygdala and brain regions involved in the DMN (21, 27), FP and VentAttn networks (27), indicating the importance of studying connectivity of the amygdala. Overall, these results indicate that most of the resting-state connectivity alterations are found between DMN, DorsAttn, VentAttn and SomMot. In contrast with findings from other pediatric psychiatric disorders, disrupted DMN-amygdala as well as DMN-FP are also observed in ADHD (28), anxiety (29) and depressive (30) disorders. Once again, these highlight the importance of clarifying the deficits in resting-state connectivity that are specifically associated with antisocial behaviors, compared to other psychiatric disorders.

Despite the relevance of the above-mentioned findings, there are several limitations that tamper scientific progress in the field. First, there are discrepancies in results across studies which may be explained by different methodologies such as restricting analyses to *a priori* defined seeds (e.g., amygdala) or a limited number of large-scale networks (e.g., DMN). Likewise, the diversity of psychometric scales used to assess antisocial behaviors and CP may contribute to heterogeneity of results. Furthermore, studies on resting-state functional connectivity usually include small sample sizes (median: 22 subjects (19)), which may increase the false positive rate. In fact, some authors have recently found that in resting-state functional connectivity investigations, stability and reproducibility in brain-behavior relationships may require thousands of individuals (31). As an alternative, authors from a recent IMAGEN study have aggregated several measures of structural brain deficits (e.g., cortical surface area, cortical thickness and subcortical volumes), similarly as in the case of polygenic risk scores, to study mental disorders in adolescents (32). They showed that a neuroimaging association score may improve reproducibility and generalizability in brain-behavior relationships compared to individual measures. To our knowledge, no studies have aimed to produce a brain connectivity score that may be generalizable regarding antisocial behaviors.

The purpose of the study was twofold. First, we aimed to address these issues by investigating the cortico-cortical and amygdala-cortical functional connectivity at rest associated with two distinct measures of CP, using a large sample of 1416 children and adolescents. We hypothesized that CP will be associated with disrupted functional connectivity within-DMN regions, and between DMN and DorsAttn, VentAttn, SomMot networks, as well as between the amygdala and these networks. Second, we sought to produce an aggregated brain connectivity score that may be specific to CD and generalizable across CP-related psychopathologies such as variants of psychopathy. To do so, we compared the aggregated connectivity score between variants of psychopathy and healthy subjects and examined the ability of the score to discriminate between CD and other pediatric psychiatric disorders (e.g., ADHD, anxiety and depressive disorders). We expected that the brain connectivity scores will differ between variants of psychopathy and healthy controls and be specifically associated with CD/ODD but not with other psychiatric disorders.

## 2. METHODS

### 2.1. Participants and Neuroimaging Acquisition Parameters

Data from 2200 participants were obtained from the Healthy Brain Network (HBN), an ongoing initiative in New York area (USA) that aims to investigate heterogeneity and impairment in developmental psychopathology (5-21 years old) (33). The HBN adopted a community-referred recruitment model in which advertisements was provided to community members, educators, parents. Exclusion criteria were impairments that prevents full participation in the study (e.g., serious neurological disorders, hearing or visual impairments), neurodegenerative disorder, acute encephalopathy, acute intoxication, and serious psychiatric disorders (recent diagnosis of schizophrenia and/or manic episode). Supplemental information is provided elsewhere (33).

From the 2200 participants included in the Data Release 7.0, 1583 participants contained available functional neuroimaging data. Written assent was obtained from participants younger than 18 years old, and written consent was obtained from their legal guardians. Written informed consent was obtained from participants aged 18 or older prior to enrolling in the study. The original HBN study was approved by the Chesapeake Institutional Review Board (https://www.chesapeakeirb.com/). The current study was approved by the local ethics committee.

MRI acquisition took place at three different sites: mobile 1.5T Siemens Avanto in Staten Island, 3T Siemens Tim Trio at Rutgers University Brain Imaging Center (RUBIC), and 3T Siemens Prisma at the CitiGroup Cornell Brain Imaging Center (CBIC) (acquisition protocols and parameters can be found in Table S1, in (33) as well as http://fcon_1000.projects.nitrc.org/indi/cmi_healthy_brain_network/). Data at the CBIC were obtained using the same data acquisition protocol implemented at RUBIC. The acquisition of the two resting-state scans lasted 5 min each, during which participants viewed a fixation cross located at the center of the computer screen. Data for the Siemens Avanto were acquired in a single run lasting 10 minutes.

### 2.2. Main Assessments

Conduct problems were assessed using the Child Behavior Checklist (CP-CBCL, (34)), which comprised 33 items from Aggressive (20 items) and Rule-Breaking (11 items) syndromes scales. Parents rated each item using a 3-point scale (0=not true to 2=very true)(α=.93). We also used the 5-item CP scale (2 items on aggressive and 3 on non-aggressive rule-breaking behaviors) of the Strength and Difficulties Questionnaire (CP-SDQ, (35)), which showed acceptable internal consistency (α=.72). Pearson’s correlation between these two scales of CP revealed moderate-strong association (*r*=.788).

### 2.3. fMRI data preprocessing

Functional images were realigned, corrected for motion artifacts with the Artifact Detection Tool (36)(ART, setting a threshold of 0.9□mm subject ART’s composite motion and a global signal threshold of Z□=□5) with the implemented in CONN Toolbox (37), bandpass filtered (0.01□Hz < f < 0.10 Hz) and co-registered to the corresponding anatomical image. The anatomical images were segmented (into GM, white matter, and cerebrospinal fluid) and normalized to the Montreal Neurological Institute (MNI) stereotaxic space. Functional images were then normalized based on structural data, spatially smoothed with a 6□mm full-width-at-half-maximum (FWHM) 3D isotropic Gaussian kernel and resampled to 2□mm^3^ voxels. For the preprocessing, the anatomical component-based noise correction method (aCompCor strategy, (38)), was employed to remove confounding effects from the BOLD time series, such as the physiological noise originating from the white matter and cerebrospinal fluid. This method was found to increase the validity and sensitivity of analyses (39). In the current study, preprocessing issues were found in 108 participants (n=1475), and 59 adolescents exhibited high movements (exceeding 3mm), leaving a final sample size of 1416 adolescents.

### 2.4. Cortico-cortical and Amygdalo-cortical functional connectivity

To examine the cortico-cortical connectivity, we used the 333 cortical parcels grouped into 13 intrinsic networks, derived from the Gordon Atlas which includes 13 intrinsic networks (15) (i.e., Auditory, Cingulo-Opercular, Cingulo-Parietal, DMN, DorsAttn, FP, VentAttn, SomMot-Hand, SomMot-Mouth, Retrosplenial-Temporal, Salience, Visual, None). We additionally included left and right amygdala from the FSL Harvard-Oxford Atlas, provided in the CONN Toolbox. Physiological noise, realignment parameters, and movement artifacts were regressed out as confounding effects from the BOLD time-series for each parcel.

In the first-level analysis, Pearson’s correlation coefficients between the residual BOLD time course from each parcel and the time course of all other 332 parcels, for each subject. The same was done for amygdala regions and all Gordon 333 parcels. Coefficients were converted to normally distributed z-scores using a Fisher Z-Transformation. Second-level analyses were conducted using mass univariate linear regression to examine relationships with CP derived from the CBCL and the SDQ, removing the effect of age, sites, sex, percentage of valid scans and framewise displacement. We ran univariate linear regression analyses on 5,000 random subsamples using 90% of the total sample at each iteration. Thresholding was performed in two steps to adequately control for both type II and type I errors, successively. First, Brain connectivity was considered as statistically associated with CP if the average p-value across the 5,000 iterations met the uncorrected threshold of p<0.005. This somewhat liberal threshold was used to keep brain connectivity that has acceptable association with CP. We then thresholded these brain connectivity by keep those that intersected between the CBCL and the SDQ to control for type I errors. Furthermore, we investigated whether brain connectivity results differed between scales regarding their importance in explaining variance of the CBCL and SDQ. Permutation importance was calculated by conducting a multivariate linear regression which included the resulting brain connectivity measures (independent variable) in association with CP severity (dependent variable), respectively. We permutated each brain connectivity measure 100 times on a test set (20% of the data) and compared R_2_ scores between the baseline model on the train set (80% of the data without permutations) and the test set 1,000 times using Monte-Carlo cross-validation. Compared to the base model, changes in R_2_ score would therefore indicate the relative importance of a particular feature. Finally, we compared the feature importance of each brain connectivity between the CBCL and SDQ with Fisher r-to-z transformation (p<0.05, two-tailed).

### 2.5. Functional Decoding

Functional decoding was conducted to examine the neurocognitive domains (i.e., task fMRI) underlying functional connectivity between two ROIs that are associated with CP. Briefly, each parcel was characterized by a binary set of behavioral categories (z > 3, p<0.05 Bonferroni correction for multiple comparisons) derived from the BrainMap database (see Behavioral Analysis Plugin for Multi-image Analysis GUI (40) (ric.uthscsa.edu/mango). In the current study, 49 categories from 4 different neurocognitive domains were included (i.e., Action, Emotion, Cognition, Perception). For each pair of brain connectivity (e.g., parcel A and parcel B), we created an adjacency matrix (49 categories-by-49 categories) representing the connected categories between parcel A and parcel B. Then, we summed these adjacency matrices for positive (i.e., 10) and negative (i.e., 11) brain connectivities associated with CP, separately. Finally, behavioral categories at a node-level were ranked based on their number of edges (degree centrality) and influence across the network (betweenness centrality). We also examined what behavioral domains were the most frequently reported across brain connectivities. These analyses were conducted with python’s NetworkX package (41)

### 2.6. Deriving an Aggregated Measure of Brain Connectivity associated with CP

In a recent IMAGEN study, the authors have aggregated measures of structural brain deficits (e.g., cortical surface, cortical thickness and subcortical volumes) and showed that the neuroimaging association score may increase reliability and generalizability in the analyses on brain-behavior relationships, as compared to individual measures (32).

In our study, we computed a neuroimaging association score by aggregating measures of functional connectivity that intersected between the CBCL and the SDQ. The brain connectivity scores were computed for both positive and negative association with CP, separately. In contrast to Axelrud and colleagues (32) in which they weighted structural measures with effect size found in ENIGMA studies, we rather summed brain connectivity measures given the absence of ROI-to-ROI functional connectivity meta-analysis in antisocial subjects. These sums of brain connectivity measures were then tested on CBCL and SDQ, separately, to test the variations between scales. We conducted univariate and multivariate linear regression analyses adjusting for age, sites, sex, percentage of valid scans and framewise displacement and other psychological confounders that are known to be related to CP such as callous-unemotional traits, irritability, anxiety, and hyperactivity/impulsivity traits.

### 2.7. Validation of the Connectivity Scores with Psychopathy Variants & Clinical Diagnoses

We further aimed to test whether the CP brain connectivity scores can be used to differentiate between variants of psychopathy and clinical diagnoses. First, variants of psychopathy in adolescents were extracted by conducting latent profile analysis (LPA) on callousness and anxiety traits (Dugré and Potvin, in revision, see Supplementary Material for LPA method). Briefly, LPA analyses revealed 4 homogenous groups: anxious adolescents, typically developing (TD), primary variant of psychopathy and secondary variant of psychopathy. In the current study, we compared brain connectivity scores between both variants and TD group by conducting a one-way analysis of variance with Tukey’s posthoc test. Compared to the TD group, the primary and the secondary variants showed higher risk of CP on CBCL (OR=1.25 & 1.27, respectively) and SDQ (OR=2.41 & 2.24, respectively).

Second, a computerized web-based version of the Schedule for Affective Disorders and Schizophrenia—Children’s version (KSADS, (42)) as administered to all participants by a licensed clinician (33). The KSADS-COMP includes a clinician-conducted parent interview and child interview, which results in automated diagnoses. In the current study, we used the consensus clinical diagnoses (i.e., presence or absence) generated by the Healthy Brain Network Team after reviewing interviews and materials for each participant (33). Diagnosis categories include Conduct Disorder/Oppositional Defiant Disorder (CD/ODD), Disruptive Mood Dysregulation Disorder (DMDD), Attention-Deficit/Hyperactivity Disorder (ADHD), Anxiety Disorders (ANX), Depressive Disorders (DEP), Autism Spectrum Disorder (ASD), and other Neurodevelopmental Disorders. Associations between brain connectivity scales and clinical diagnoses were conducted by examining feature importance, similarly as above. In a multivariate logistic regression analysis, we examined the Odds Ratios (OR) and significance of positive and negative brain connectivity scores associated with clinical diagnoses, while adjusting for covariates (i.e., age, sites, sex, percentage of valid scans and framewise displacement). Finally, we investigated the ability to discriminate diagnosed subjects from healthy controls, using only the brain connectivity scores (predicted probabilities), and plotted the Area Under the Receiver Operating Characteristic (ROC) curve (43).

## 3. RESULTS

### 3.1. Cortico-cortical and Amygdalo-cortical functional connectivity

Mass univariate functional connectivity unveiled significant connectivities associated with CP-CBCL (231 connections) and CP-SDQ (269 connections). However, only 21 connections intersected between the two scales (10 positive and 11 negative associations with CP, see Table 1 & Figure 1A-C). Overall, CP was mainly associated with the somatomotor (6 out of 21 connections) and ventral attention networks (4 out of 10 positive connections), but also with unassigned parcels from the Gordon Atlas (None: 4 connections). More precisely, severity of CP was positively associated with functional connectivity within-SomMot (2 connections), between FP and unassigned parcels (i.e., bilateral pHippocampus-FEF) but also between VentAttn and DMN, DorsAttn, FP and SomMot. Furthermore, CP was negatively associated with functional connectivity between cingulo-opercular & Visual (2 connections), SomMot & Salience network (2 precentral-dACC), auditory & cingulo-opercular & DMN as well as within-DMN.

**Table 1.** Significant Results in resting-state connectivity intersecting between the CBCL & SDQ

| Connectivity |  |  | Statistics |  |  |  |
| --- | --- | --- | --- | --- | --- | --- |
|  |  |  | CP-CBCL |  | CP-SDQ |  |
| Name | Parcel 1 (Network) | Parcel 2 (Network) | F | *p* | F | *p* |
| *Positive Associations* |  |  |  |  |  |  |
| P1 | pSMG (DA #24) | TPJ (VA #12) | 9.32 | 0.004 | 9.69 | 0.003 |
| **P2** | **Lateral PFC (FP #4)** | **vlPFC (VA #8)** | **14.12** | **<0.001** | **9.21** | **0.004** |
| P3 | pHippocampus (None #23) | Frontal Eye Field (FP #24) | 10.45 | 0.002 | 9.44 | 0.004 |
| P4 | pHippocampus (None #3) | Frontal Eye Field (FP #24) | 14.57 | <0.001 | 12.25 | 0.001 |
| P5 | Postcentral (SH #16) | Precentral (SH #29) | 10.65 | 0.002 | 9.34 | 0.004 |
| P6 | Postcentral (SH #18) | Precentral (SH #29) | 8.89 | 0.005 | 10.15 | 0.003 |
| P7 | Precentral (SH #23) | pITG (None #40) | 9.34 | 0.004 | 9.07 | 0.004 |
| **P8** | **Premotor (SH #30)** | **lateral OFC (VA #21)** | **9.33** | **0.004** | **9.22** | **0.004** |
| P9 | Precentral (SM #2) | dlPFC (DA #8) | 9.01 | 0.004 | 12.56 | 0.001 |
| P10 | Angular (VA #2) | Precuneus (DMN #28) | 11.24 | 0.001 | 12.87 | 0.001 |
| *Negative Associations* |  |  |  |  |  |  |
| N1 | pINS (A #9) | dmPFC (D #39) | 13.48 | 0.001 | 9.51 | 0.004 |
| N2 | aINS (CO #37) | Lingual (V #37) | 13.53 | <0.001 | 12.57 | 0.001 |
| N3 | SMA (CO #5) | Heschl (A #23) | 10.46 | 0.002 | 10.42 | 0.002 |
| **N4** | **dmPFC (D #4)** | **Lateral PFC (D #33)** | **11.97** | **0.001** | **10.14** | **0.003** |
| N5 | Lateral OFC (None #7) | ITG (None #35) | 10.41 | 0.002 | 9.24 | 0.004 |
| **N6** | **Lateral OFC (None #8)** | **SMA (CO #27)** | **12.22** | **0.001** | **13.01** | **0.001** |
| N7 | dACC (S #1) | Postcentral (SH #18) | 8.79 | 0.005 | 10.97 | 0.002 |
| N8 | dACC (S #1) | Postcentral (SH #37) | 8.98 | 0.004 | 11.80 | 0.001 |
| N9 | Precentral (SH #29) | pMTG (None #34) | 10.62 | 0.002 | 11.62 | 0.001 |
| N10 | Postcentral (SM #4) | pgACC (D #23) | 15.22 | <0.001 | 13.68 | <0.001 |
| N11 | V2 (V #17) | Lateral PFC (CO #20) | 19.26 | <0.001 | 10.46 | 0.002 |

*Note.* Names of the brain regions were derived from the center coordinates of each parcel using the Anatomy Toolbox (44). Top 5 most important features are in **BOLD**.

**Figure 1.**
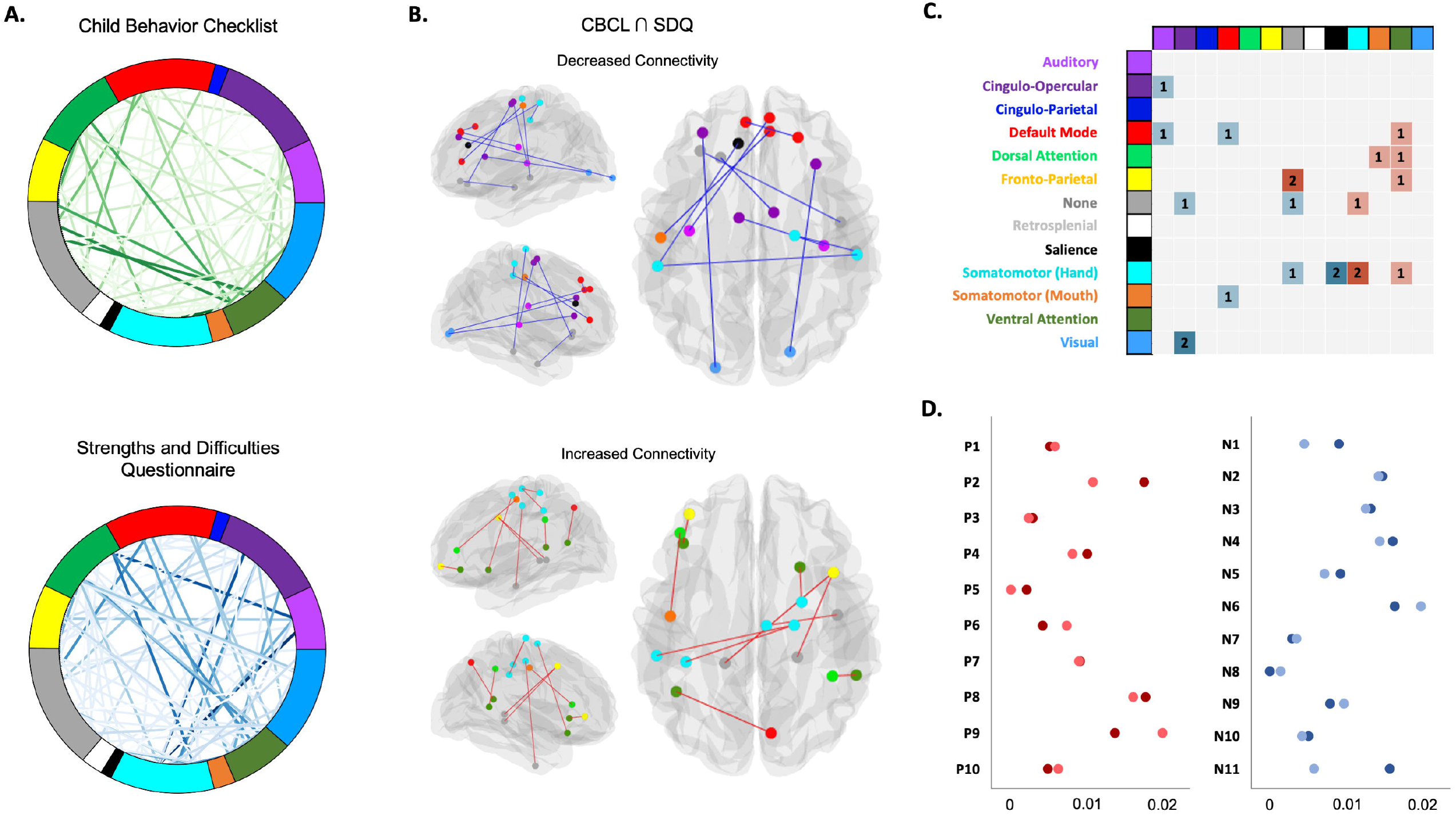
Associations between cortico-cortical connectivity and Conduct Problems across different scales. **A**. Weight (F-value) of each significant (p<0.005) cortico-cortical connectivity across 13 networks of the Gordon (333 parcels, Gordon et al., 2015) after 5,000 random subsampling using 90% of the sample in association with Conduct Problems scales derived from the Child Behavior Checklist (CP-CBCL) and the Strengths and Difficulties Questionnaire (CP-SDQ). **B**. Connectivity positively and negatively associated with Conduct Problems that intersected between the CBCL and SDQ (Red edges = positive associations; Blue edges = negative associations). **C**. Adjacency matrix showing significant within- and between-network connectivity results associated (Red=Positively; Blue=Negatively) with CP. **D**. Feature importance (R^2^ score with Standard Deviation) in association with severity of Conduct Problems for the Child Behavior Checklist (CP-CBCL) and the Strengths and Difficulties Questionnaire (CP-SDQ). Permutation importance was conducted by permutating each of the 21 cortico-cortical brain connectivity in a multivariate linear regression 100 times on a test set (20% of the data) repeated 1,000 using Monte-Carlo cross-validation. Red dots=brain connectivity positively associated with CP; Blue dots=brain connectivity negatively associated with CP. Darker colors=CBCL & Lighter colors=SDQ. Please refer to Table 1. for more detailed information about brain connectivity.

When examining feature importance of the 21 connections in a multivariate linear regression, we observed that between CBCL and SDQ, brain connectivity measures had relatively similar importance (see Supplementary Material for full list). The top 5 most important features were (please refer to Table 1): 1) **P8 connection**: Premotor-Lateral OFC (R_2_ change .016-.018); 2) **N6 connection**: Lateral OFC-SMA (R_2_ change .016-.020); 3) **P9 connection:** Precentral-dlPFC (R_2_ change .014-.020); 4) **P2 connection:** Lateral PFC-vlPFC (R_2_ change .011-.018) and 5) **N4 connection**: dmPFC-Lateral PFC (R_2_ change .014-.016). The least important feature was N8 connection: dACC-Postcentral (R_2_ change 0-.001).

Regarding amygdalo-cortical functional brain connectivity, analyses revealed that the CP-CBCL was negatively associated with functional connectivity between the right amygdala and the left (F= 10.38, p= 0.002) and right (F= 9.16p= 0.004) ventral PCC (BA 23). Additionally, the CP-SDQ only showed negative association between the right amygdala and the pMTG (FP) (F=12.57, p<0.001 uncorrected). Thus, no significant connectivity intersected between the two scales, which suggest low reliability in amygdala connectivity across CP scales.

### 3.2. Functional Decoding

As shown in Figure 2., the functional brain connectivity measures associated with CP were characterized by a variety of behavioral categories. First, positive brain connectivity measures were mainly related to interaction between Action and Cognition as well as within-Cognition domains. Indeed,, the most frequent connection of behavioral domains was between Speech Execution (Action) and Working Memory (Cognition) with 4 out of 11 pairs of parcels. Also, the top 5 categories with the largest number of connections (node centrality) included: Unspecified (Action), Speech Execution (Action), Working Memory (Cognition), Attention (Cognition) and Semantics (Cognition). The top 5 categories that had the most influence (betweenness centrality) on the network were: Working Memory (Cognition), Unspecified (Action), Speech Execution (Action), Explicit Memory (Cognition) and Semantics (Cognition).

**Figure 2.**
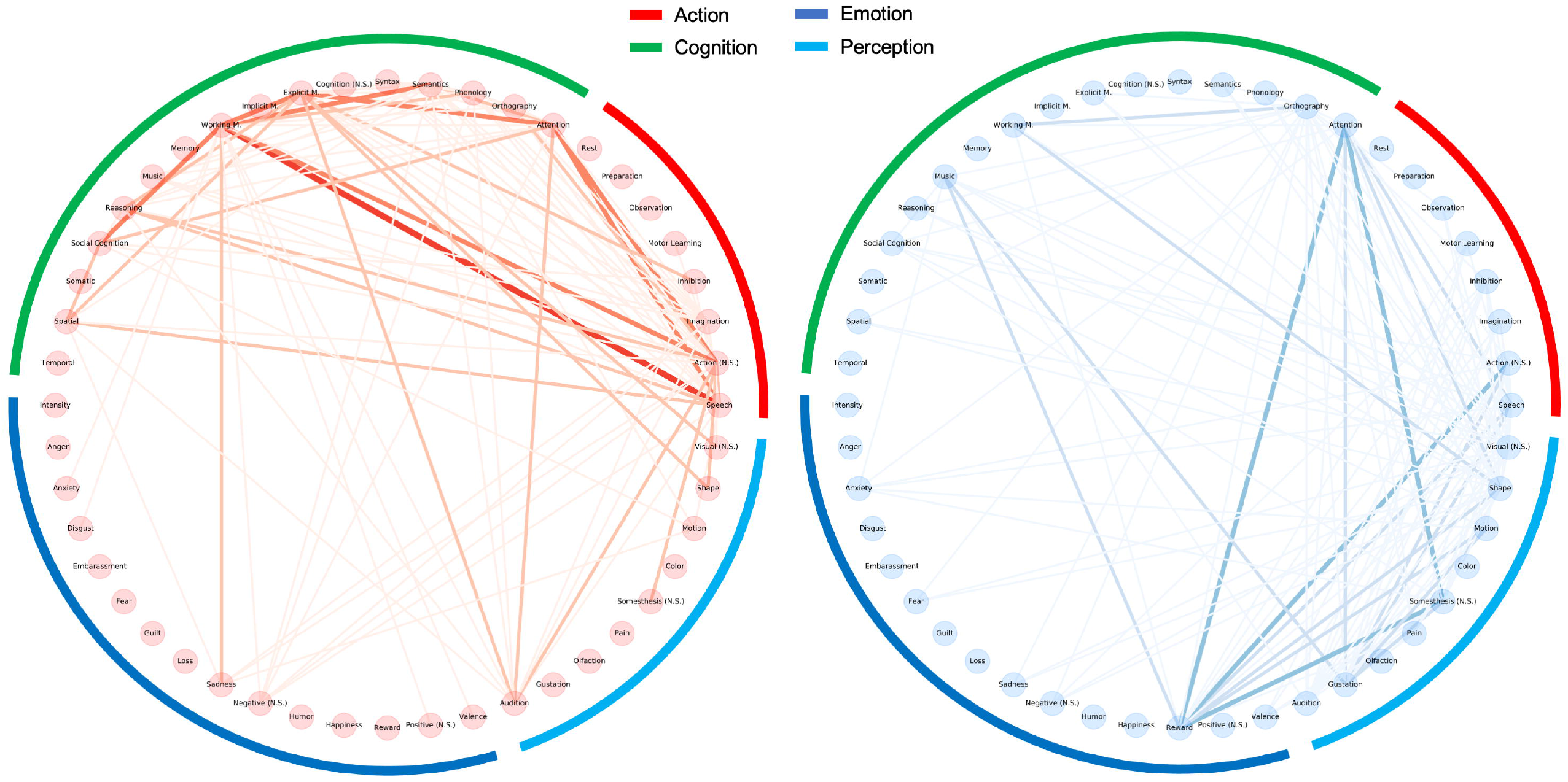
Circular layout displaying the relationship between the behavioral domains significantly associated with functional brain connectivity. Red graph = brain connectivity positively related to CP. Blue Graph = brain connectivity negatively related to CP. Thicker line represents larger number of connected behavioral categories across pairs of brain connectivity.

Second, negative brain connectivity measures rather showed a widespread relationship between the four behavioral domains. Indeed, the most frequent connections were 1) Action Execution (Unspecified) & Reward, 2) Attention & Reward, 3) Attention & Somesthesis (Unspecified), and 4) Reward & Somesthesis (Unspecified) with each 3 out of 10 pairs of parcels (Figure 2). Moreover, the top 5 categories with the largest number of connections (node centrality) were: Orthography (Cognition), Shape (Visual), Unspecified (Visual), Speech Execution (Action) and Attention (Cognition). Finally, the top 5 categories that had the most influence on the network were: Orthography (Cognition), Shape (Visual), Unspecified (Visual), Speech Execution (Action) but also Reward (Emotions).

### 3.3. Deriving an Aggregated Measure of Brain Connectivity

We subsequently aggregated the brain connectivity measures associated with CP for both positive and negative associations, separately and investigated whether the relationships between these scores and conduct problems may vary across the two different scales. Regarding the CBCL, univariate linear regression revealed significant association between both positive (*B*=3.29, p<0.001, R_2_=.051 and negative (*B*=-3.65, p<0.001, R_2_=.06) connectivity scores and CP-CBCL. After adjusting for sociodemographic and clinical covariates both the positive (*B*=1.20, p<0.001), and negative (*B*=-1.44, p<0.001) brain connectivity scores remained significantly associated with CP. This was irrespective of subfactors of CP such as aggressive and rule-breaking behaviors. In fact, positive brain connectivity score was associated with both aggressive (*B*=1.14, p<0.001) and RB (*B*=.84, p=0.002) subfactors of CP. This was also found for negative brain connectivity score in relationship with aggressive (*B*=-1.28, p<0.001) and RB (*B*=-1.18, p<0.001) behavior scores.

For SDQ, univariate linear regression revealed significant association between both the total score of positive (*B*=.63, p<0.001, R_2_=.047) and negative (*B*=-.73, p<0.001, R_2_=.054) connectivity and CP-SDQ. Both positive (*B*=.33, p<0.001), and negative (*B*=-.29, p<0.001) brain connectivity scores remained statistically significant after adjusting for covariates.

### 3.4. Validation of the Connectivity Scores with Psychopathy Subtypes & Clinical Diagnoses

First, we then investigated whether these brain connectivity scores differ between variants of psychopathy and healthy adolescents (Figure 3A). Analysis of variance revealed that positive (F_(2, 334)_=13.46; p<0.001) and negative (F_(2, 334)_=3.45; p=0.03) scores significantly differed between groups. Posthoc tests showed that both variants showed higher positive scores than healthy adolescents (p<0.001), whereas only the primary variant demonstrated lower negative score than healthy adolescents (p=0.035).

**Figure 3.**
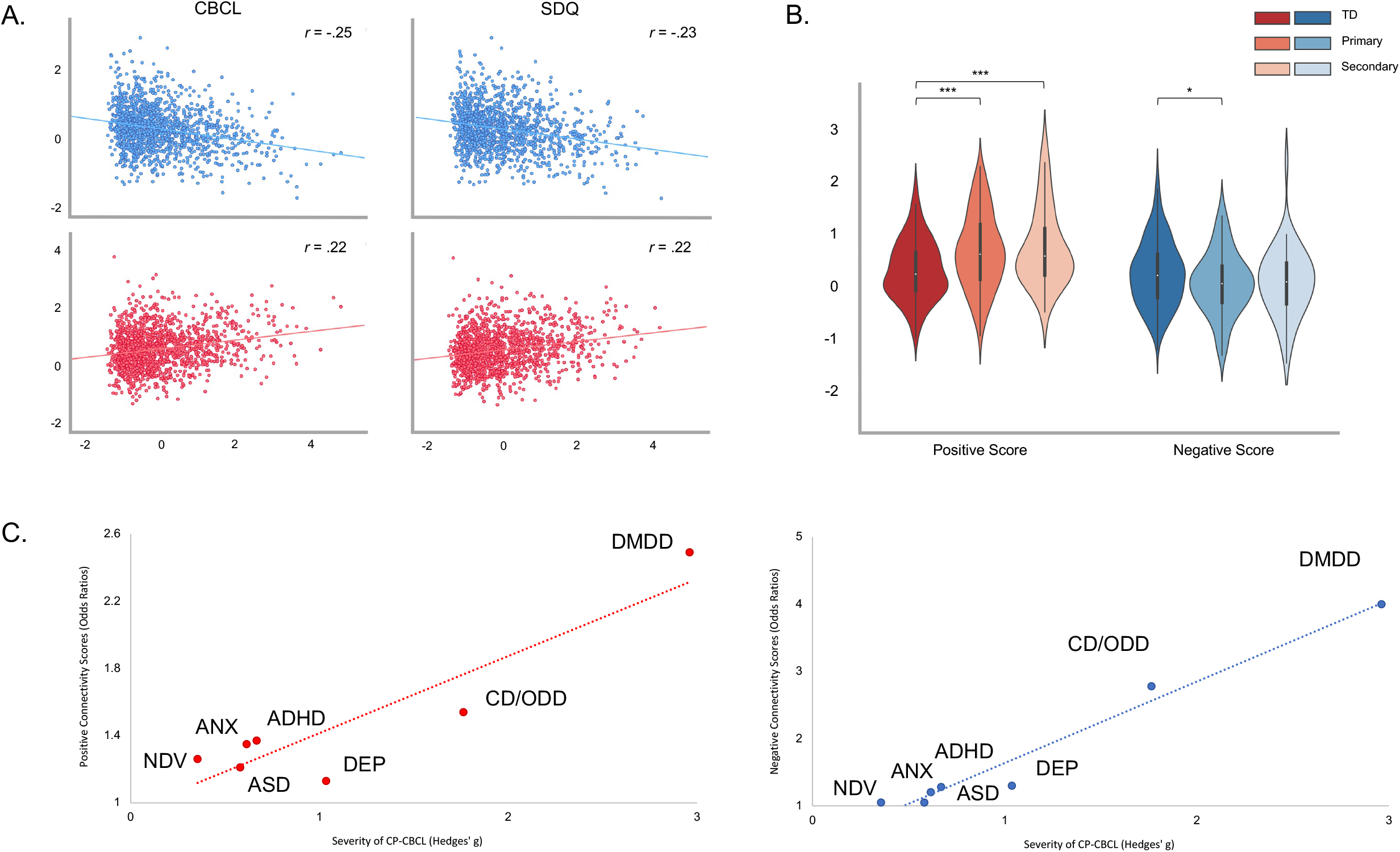
Results from the Brain Connectivity scores and their external validation. **A**. These scatter plots represent the relationship between positive (red) and negative (blue) connectivity scores in associated with CBCL and SDQ conduct problems subscales. **B**. Differences between TD and variants of psychopathy (Primary & Secondary) on positive and negative brain connectivity scores *p<0.05; **p<0.01; p<0.001. **C**. Association between brain connectivity scores (Odds ratios) and severity of conduct problems (Hedge’s g compared to healthy controls) for each psychiatric disorder. For the negative brain connectivity score we computed the inverse Odds Ratios (1/OR).

Second, while adjusting for covariates, multivariate logistic regression models showed (Figure 3B) that both brain connectivity scores significantly increased the risk for DMDD, CD/ODD (only negative connectivity score), ADHD and ANX (only positive connectivity score) compared to subjects without any psychiatric disorder. These associations remained statistically significant (except for ANX) when examining the risk of a given diagnosis versus others (i.e., healthy controls and other diagnosis) (see Supplementary Material). Using only the brain connectivity scores, analyses revealed good to acceptable ability to discriminate DMDD (AUC=.70, 95% CI: .60-.80) and CD/ODD (AUC=.65, 95% CI: .57-.73), but poor classification ability for ADHD (AUC=.58, 95% CI:.54-.62) (Figure 3C). These results are supported by the fact that DMDD (Hedges’s g=2.46-2.96), CD/ODD (Hedges’s g=1.67-1.76), and ADHD to a lesser extent (Hedges’s g=0.55-0.67) displayed higher levels of antisocial behaviors (CBCL and SDQ) compared to healthy controls (see Figure 3C). Furthermore, we observed that the brain connectivity scores (Odds ratios) were closely associated with severity of CP (Hedges’s g in comparison to healthy controls) across psychiatric disorders (Figure 3C).

## 4. DISCUSSION

Using a large sample of adolescents, we aimed to clarify the role of cortico-cortical and amygdalo-cortical functional brain connectivity associated with CP. More precisely, we investigated the reliability of the relationship between resting-state brain connectivity measures and severity CP using two different psychometric scales (CBCL and SDQ). We observed that only 21 cortico-cortical (but no amygdalo-cortical) resting-state connectivity measures associated with CP overlapped between the two scales (10 positive and 11 negative associations). These brain connectivities mainly included parcels from the SomMot, VentAttn and FP networks (positive associations) as well as Cingulo-Opercular, Salience and DMN regions (negative associations). Additional analyses revealed that these regions were characterized by interactions between Action & Cognition (i.e., Positive association with CP) as well as between Reward and Cognition, Perception and Action (i.e., Negative associations with CP). Furthermore, we sought to produce brain connectivity scores that may be generalizable, by aggregating brain connectivity measures. We observed that positive and negative brain connectivity scores were reliably related to CP, with small variations between the two scales. More importantly, these brain connectivity scores differed between variants of psychopathy and healthy subjects and showed specificity for CD/ODD and DMDD. These suggests that the connectivity results may be reliable and useful estimate in our understanding of the neurobiological markers of CP.

In our recent meta-analysis of resting-state connectivity studies, we showed that antisocial subjects exhibited hyperconnectivity with ventral attention network (ie., aMCC/pre-SMA) and amygdala, and hypoconnectivity regions of the DMN (i.e., mPFC and PCC/Precuneus) and Dorsal attention network (i.e., PMC, SPL), compared to healthy controls (45). In line with these results, we found that CP was positively associated with 4 brain connectivity including regions of the ventral attention network and negatively associated with 2 brain connectivity that involved parcels of the DMN. However, contrasting with results from the meta-analysis, we found that CP was rather prominently associated with disrupted connectivity from the SomMot network (7 connections), from brain regions unassigned to any of the Gordon Networks such as posterior hippocampus and inferior/middle temporal gyri (6 connections), FP (3 connections) as well as cingulo-opercular networks (3 connections). Moreover, we found no reliable evidence of amygdala-cortical connectivity across scales. It is noteworthy to mention that studies included in our prior meta-analysis restricted their analyses on *a priori* seeds and did not investigate the whole connectome, and this may explain the discrepancies between results. Also, the functional connectivity alterations associated with CP may differ between a case-control design versus a study examining severity of CP, dimensionally. For instance, dimensional analysis (whole-brain) reveals deficient amygdala activation across task-based fMRI studies, but not case-control design (10). In contrast, case-control analysis yields disrupted amygdala resting-state connectivity, but not dimensional analyses (19). Disentangling differences in amygdala deficits between dimensional and case-control designs in relationship with CP should be prioritized in future studies given the emphasis of this brain region in the pathophysiology of CD.

Interestingly, we provided evidence that brain connectivity measures that were positively associated with CP were mainly characterized by interaction between Action Execution & Cognition behavioral domains, whereas those negatively correlated with CP were mostly represented by interactions between Reward and Action Execution, Attention & Somesthesis. First, brain connectivity measures that were positively associated with CP included lateral PFC regions (i.e., ventro and dorsolateral) and the postcentral/precentral gyri. According to a recent meta-analysis, both the lateral PFC and precentral gyrus co-activate during n-back working memory tasks (46). Indeed, it has been shown that the lateral PFC (ventral and dorsal parts) plays a major role in the reception, maintenance and monitoring of sensory inputs and sending outputs to the motor system (47, 48), whereas the precentral gyrus may rather be involved in action preparation and the processing of motor movements (49). As such, these results are in line with a recent meta-analysis of task-based fMRI studies showing that antisocial subjects exhibit aberrant co-activation of these particular brain regions (i.e., precentral and ventrolateral prefrontal cortex) during cognitive control tasks (10). Second, brain connectivity measures that were negatively associated with CP mainly included the pg-& dACC, the SMA and the aINS and lateral PFC. In contrast with the functional decoding suggesting their implications in reward processing tasks, we recently found that antisocial subjects exhibited reduced response in these regions (i.e., pg-& dACC extending to the aMCC/pre-SMA as well as the aINS) during acute threat response (10). While they are systematically observed across meta-analyses on reward tasks (50-54), the ACC and aINS are not specific to any particular neurocognitive domain (55, 56). In fact, they are known to be involved in detecting behaviorally relevant stimuli in the environment (17), in general, and may play an interacting role between internally (i.e., DMN) and externally directed actions (i.e., frontoparietal network) (57). More importantly, during a reward-interference task, some authors showed that the pg-& dACC, MCC, SMA and insula/lateral PFC were all negatively correlated with severity of instrumental motivation to aggress their opponent (e.g., ‘*in order to win*’) (58). Despite that there are very few fMRI studies that aimed to examine brain correlates of proactive aggression, these nonetheless concurs with our findings. Further research should explore this more specifically.

Finally, we demonstrated the usefulness of aggregating brain connectivity measures across scales by showing that variants of psychopathy, irrespectively of severity of CP, significantly differed from healthy controls on both scores (positive and negative). Indeed, as described earlier, positive and negative brain connectivity scores may be related to cognitive control and detecting relevant stimuli to adequately select behavioral response, respectively. These results concur with past studies suggesting that both variants exhibit higher impulsivity compared to healthy controls (59-62), although some have found that the secondary variant may display poorer impulse control than the primary variant (61, 63). Additionally, adolescents with the primary variant are thought to be prone to proactive aggression and thrill and novelty seeking (59, 60, 64), compared to their counterparts with the secondary variants, which may explain that the negative brain connectivity score did not differ between secondary variant and healthy controls. Furthermore, we showed that both brain connectivity scores significantly increased the risk for DMDD, CD/ODD (except for the positive brain connectivity score) and ADHD. As the positive brain connectivity score mainly include SomMot regions, this concurs with past evidence indicating that deficient activity of the Presupplementary motor area and precentral gyrus during inhibition conferred an increased risk for general externalizing behavior (i.e., ADHD, Conduct Disorder and Substance Misuse) (65).

Likewise, negative brain connectivity score was mainly represented by brain regions of the Salience, VentAttn and Cingulo-Opercular networks such as the dACC and bilateral vlPFC and aINS. Dysconnectivity in these networks were found to be associated with the broad dimension of externalizing pathology in two recent studies ((66) and ABCD study:(67)) which corroborates our findings.

## LIMITATIONS

In our study, we aimed to address several limitations of current literature on resting-state functional connectivity, by using a large sample size of adolescents. However, a few limitations need to be acknowledged. Indeed, the sample contains a relatively wide age range spanning from childhood to late adolescence. Although this could have introduced biases in results, we took additional measures to minimize the effects of age. Indeed, age did not alter the relationship between brain connectivity scores and CP. Secondly, neuroimaging data was collected in 3 different sites that may have altered results. It should be noted, however, that two sites used similar scanning parameters, and that we also tested the effects of sites on our results to examine the potential biases. Third, the HBN adopted a community-referred recruitment model. Therefore, careful interpretations should be made when comparing study results with population-based cohorts. Finally, we did not have a validation dataset to replicate our findings. However, we are confident that our results may be generalizable given the large sample size and the method used to extract brain connectivity measures.

## CONCLUSIONS

In conclusion, we found that brain connectivity associated with CP largely depends on the measure used. In fact, only 21 connections were shared between the CBCL and SDQ even if they strongly correlated at .79. Nonetheless, these 21 connections mainly spanned the SomMot, VentAttn and FP (positive) and Cingulo-Opercular, Salience and DMN (negative) networks. Furthermore, the aggregated scores significantly distinguished variants of psychopathy as well a pediatric psychiatric disorder, indicating reliable estimates of CP. Thus, future studies should aim to replicate our results in order to increase our understanding of the neurobiological markers of CP.

## Data Availability

All data produced in the present study are available upon reasonable request to the authors

## Acknowledgements

JRD is holder of a PhD scholarship from the Fonds de Recherche du Québec en Santé. SP is holder of the Eli Lilly Canada Chair on schizophrenia research.

